# Combined Effect of Inflammation and Malnutrition for Long-term Prognosis in Patients with Acute Coronary Syndrome: A Cohort Study

**DOI:** 10.1101/2023.12.11.23299837

**Authors:** Yang Yuxiu, Xiaoteng Ma, Fei Gao, Tao Liu, Jianping Deng, Qiuxuan Li, Zaiqiang Liu, Yufei Wang, Yang Zheng, Jingyao Yang, Zhijian Wang

## Abstract

**Background:** Dysregulated inflammation with superimposed malnutrition may constitute a significant threat in acute coronary syndrome (ACS), which needs to be elucidated. We aimed to explore the prevalence and combined predictive value of inflammation and malnutrition in ACS patients.

**Methods:** Patients presenting with ACS undergoing percutaneous coronary intervention (PCI) were retrospectively included and stratified into four groups: nourished without elevated high-sensitivity C-reactive protein (hs-CRP), nourished with elevated hs-CRP, malnourished without elevated hs-CRP, and malnourished with elevated hs-CRP. Elevated hs-CRP was determined as over 2.2mg/L, and malnutrition was defined according to the nutritional risk index (NRI). The primary outcome was major adverse cardiovascular events (MACEs), the composite of cardiac mortality, non-fatal myocardial infarction, non-fatal stroke, and unplanned revascularization.

**Results:** A total of 1,743 patients were ultimately included; 646 (37%) presented elevated hs-CRP, and 119 (7%) were considered malnourished. During a median follow-up of 30 months, 351 (20.1%) MACEs occurred. The coexistence of malnutrition and elevated hs-CRP correlated with the worst outcomes among the four phenotypes, with a significantly increased risk of MACEs (adjusted hazard ratio: 2.446; 95% confidence interval: 1.464 - 4.089; p <0.001). In the subgroup analysis, NRI displayed MACEs-predicting value merely among patients with elevated hs-CRP rather than those without (p for interaction = 0.005), suggesting the modifying effect of inflammation; simultaneously, the prognostic implications of hs-CRP were influenced by patients’ baseline nutritional status, as it only existed in malnourished patients (p for interaction < 0.001).

**Conclusions:** Among patients with ACS undergoing PCI, the double burden of inflammation and malnutrition signifies poorer outcomes, and their prognostic implications may be amplified by each other, which would provide implications to facilitate more individualized ACS care.

## Introduction

Acute coronary syndrome (ACS) is still among the leading causes of global mortality and morbidity regardless of region or ethnicity, even though there has been substantial scientific progress and mature management systems^1^. As a systemic and dynamic process, inflammation plays multiple maladaptive roles contributing to the progression and destabilization of ACS, including atherogenesis, plaque evolution, thrombogenesis, as well as myocardial damage and repair ^2,3^. Decades of evidence have accumulated supporting that using high-sensitivity C-reactive protein (hs-CRP), white blood cell (WBC) count, or the neutrophil-lymphocyte ratio (NLR) as an effective systemic inflammatory indicator among ACS patients to guide risk stratification and therapeutic approaches ^4–6^. However, it’s still pending to identify suitable patients who would benefit more from the proactive anti-inflammatory therapy on the road to more individualized ACS care.

Coronary artery disease (CAD) has long been addressed as one of the “diseases of affluence”, a corollary to the population-wide rise in overnutrition and increasing income in society^7^; studies in recent years have raised a particular concern that malnutrition is not insubstantial in patients with ACS in both developed and developing countries, even among those obese^8,9^. Malnutrition fosters an increased burden through the vicious cycle of sarcopenia, cardiac dysfunction, and cardiogenic cachexia in cardiovascular diseases (CVD) ^10,11^, and it’s also a modifiable risk factor on which physicians could act^12^; however, the emphasis of which has not been placed enough in the current ACS guidelines and clinical practice ^13,14^. There is no one universal screening method for malnutrition but rather several diagnosing tools that have been validated for different settings; the nutritional risk index (NRI), the controlling nutritional status (COUNT) score, and the prognostic nutritional index (PNI) has been commonly used in these years’ studies for their simplicity and accessibility, each of them presents with different discrimination abilities in diverse diseases or conditions^15,16^.

Dysregulated inflammation with superimposed undernutrition may constitute a significant yet relatively concealed threat in ACS pathophysiology, which needs additional attention and integrated responses. Here, we aimed to explore the combined effects and interplay of inflammation and malnutrition on outcomes in ACS patients to provide valuable information facilitating optimizing individualized care.

## Methods

### Study design and population

In this single-center, retrospective observational cohort, patients presenting with ACS undergoing percutaneous coronary intervention (PCI) between June 2016 and November 2017 at Beijing Anzhen Hospital were consecutively included. Patients with missing data on nutrition-inflammation stratification or a known history of coronary artery bypass grafting were excluded. Baseline demographic data, previous medical history, clinical presentations, laboratory and echocardiographic results, angiographic and procedural characteristics, and medications were collected from the electronic medical records. This study followed the Helsinki Declaration of Human Rights and was approved by the institutional review board (IRB) of Beijing Anzhen Hospital (IRB number: 2016034x). The IRB waived the need for written patient consent as this study involved a retrospective analysis of clinically acquired data.

### Definitions

Inflammatory status was preliminarily evaluated by the level of circulating hs-CRP, WBC counts, and NLR (measured by absolute neutrophil count/total lymphocyte count), and hs-CRP was finally chosen to stratify high-inflamed patients with the cut-off value of 2.2mg/L due to its highest predictive ability among the three indices; Nutritional status was graded using the NRI, COUNT score, and PNI, respectively, and the NRI was ultimately chosen as the malnutrition criterion, given its better predictive ability than COUNT or PNI in our cohort **(Supplementary Table 1)**. The NRI was calculated using [1.519 × serum albumin (g/L)] + 41.7 × (current weight/ideal weight)] for patients under the age of 65 and [1. 489×serum albumin (g/L)] + 41.7 × (current weight/ideal weight)] for those over 65, the latter formula is also known as the geriatric NRI (GNRI)^17^. The ideal body weight was considered using the Lorenz formula, that is, [height (cm) −100 - ([height (cm) - 150]/4)] for men, and [height (cm)-100-([height (cm)-150]/2.5)] for women^18^. Scores of 97.5–100, 83.5–97.5, and <83.5 were considered mild, moderate, and severe malnutrition in patients younger than 65, while patients over 65 using scores of 92–100, 82-92, and <82 to stratify mild, moderate, and severe malnutrition^17^. The CONUT score was calculated as a sum of the scores based on the serum albumin, total cholesterol concentration, and absolute lymphocyte count and classified as normal, light, moderate, or severe for scores of 0-1, 2-4, 5-8 and 9-12, respectively^19^. PNI was calculated as [serum albumin (g/l) +5 × total lymphocyte count (10^9^/L)], and scores of 35-38 and <35 reflect moderate and severe malnutrition, respectively^20^. Standard modifiable cardiovascular risk factors (SMuRFs) include hypertension, diabetes, hypercholesterolemia, and current smoking, and SMuRF-less refers to no evidence of SMuRF^21^. Anemia was defined as hemoglobin < 120 g/L for men and <110 g/L for women^22^.

### Outcomes

The primary outcome was major adverse cardiovascular events (MACEs), defined as the composite proportion of cardiac mortality, non-fatal MI, non-fatal stroke, and unplanned revascularization. The study index date was the admission date, and complete follow-up was until any MACE occurred or December 2020, whichever came first. Patients were followed up every six months by telephone or through available medical records.

### Statistical analysis

Categorical variables were summarized as frequencies and percentages; numerical variables were summarized as the mean (standard deviation, SD) or median (interquartile range, IQR), depending on the data distribution. Categorical variables were compared with the χ² test, and continuous variables with Mann-Whitney non-parametric tests. Receiver-operating characteristic curve (ROC) analyses were performed to estimate the discriminative abilities of the nutritional scores and systemic inflammatory indicators on MACEs. The associations between the variables and the binary study outcomes were assessed using multivariable Cox proportional hazards regression models with a hazard ratio (HR) and 95% confidence interval (CI) in the total population and by stratified groups. Besides, the interactions between malnutrition and inflammation were tested by conducting likelihood ratio tests. Potential confounders selected by univariable Cox hazards analysis with p <0.05 were included in adjusted models, which were heart rate, pulse pressure, family history of CAD, diabetes, peripheral artery disease (PAD), old MI (OMI), prior PCI, anemia, WBC, triglycerides (TG), total cholesterol (TC), low-density lipoprotein cholesterol (LDL-C), high-density lipoprotein cholesterol (HDL-C), B-type natriuretic peptide (BNP), CrCl, uric acid, fasting plasma glucose (FPG), glycated hemoglobin A1c (HbA1C), left ventricular systolic dysfunction (LVSD), multivessel coronary artery disease, SYNTAX score, complete revascularization, discharged with aspirin plus clopidogrel, and discharged with beta-blocker; to be clear, systemic or diastolic blood pressure, neutrophil-lymphocyte ratio, hs-CRP, and albumin were not included in the adjusted model to avoid over adjustment bias even though their significant correlation with outcomes **(Supplementary Table 2)**. Kaplan-Meier survival probability estimates were calculated from the date of PCI up to the total available follow-up at three years, stratified according to nutritional and inflammatory status, and assessed with the log-rank test. The concordance index, net reclassification improvement, and integrated discrimination improvement statistical analyses were performed to evaluate the incremental value of the hs-CRP or NRI in addition to the GRACE (Global Registry of Acute Coronary Events) risk model. All analyses were done with R software (version 3.5.0, Austria) and Prism software (version 5.0, USA). A two-sided p-value of less than 0.05 was considered to indicate statistical significance.

## Result

A total of 1,743 ACS patients treated with PCI who met the enrollment criteria were included, with an average age of 60 years, of whom 76.5% were male. Over a median follow-up of 30 months (IQR, 30–36months), 351 (20.1%) MACEs occurred, including 37 (2.1%) cardiac deaths, 43 (2.5%) non-fatal MI, 22 (1.3%) non-fatal stroke, and 249 (14.3%) unplanned coronary revascularization. The baseline characteristics of patients stratified by the occurrence of MACEs were summarized in **Supplementary Table 3**, thereinto 50%, 7%, and 0.3% of ACS patients were classified as malnourished by COUNT, NRI, and PNI, respectively, and no one was considered severely malnourished by any score.

Determined by ROC and Cox regression analyses, hs-CRP over 2.2mg/L and NRI were used to identify high-inflamed and malnourished patients, respectively **(Supplementary Table 1)**. Patients were stratified into four phenotypes according to their nutritional status and hs-CRP categories as nourished without elevated hs-CRP group (n=1051), nourished with elevated hs-CRP group (n=573), malnourished without nourished group (n=46), and malnourished with nourished group (n=73). Compared with the other groups, malnourished patients with elevated hs-CRP were more prone to complicated diabetes, anemia, and renal dysfunction, presenting with larger proportions of AMI, worse cardiac function, and more complex coronary lesions; they were also more often to be prescribed with furosemide on discharge. After 30 months of follow-up, patients in the malnourished and elevated hs-CRP group showed the highest rates of MACEs (41.1%), cardiac death (12.3%), non-fatal MI (4.1%), and unplanned revascularization (24.7%) among the four groups (**Table 1**). The adjusted impact of inflammation or malnutrition on outcomes is shown in **Table 2**. Malnutrition showed a significant correlation to the subsequent cardiac death, unplanned revascularization, and overall MACEs; the high-inflamed status was related to the increased MACEs, mainly driven by the incidence of unplanned revascularization. Kaplan-Meier survival analyses showed that malnourished patients with elevated hs-CRP levels exhibited the most unfavorable prognosis in terms of MACEs, cardiac death, and unplanned revascularization. However, no difference among the four groups was found in non-fatal MI or non-fatal stroke (**Figure 1**).

**Table 1.** Baseline clinical characteristics and outcomes of patients stratified by the nutritional and inflammatory statuses.

| Variable | Total population<br>(n=1743) | Nourished without<br>elevated hs-CRP<br>(n=1051) | Nourished with<br>elevated hs-CRP, <sup>a</sup><br>(n=573) | Malnourished without<br>elevated hs-CRP<br>(n=46) | Malnourished with<br>elevated hs-CRP <sup>a</sup><br>(n=73) | p-value |
| --- | --- | --- | --- | --- | --- | --- |
| Demography and anthropometric data |  |  |  |  |  |  |
| Age, years | 60 (53, 67) | 61 (53, 66) | 61 (52, 68) | 63 (58, 69) | 62 (54, 73) | 0.028 |
| Sex, male, n (%) | 1333 (76.5) | 824 (78.4) | 428 (74.7) | 29 (63.0) | 52 (71.2) | 0.030 |
| Body mass index, kg/m<sup>2</sup> | 25.2 (23.6, 27.6) | 25.3 (23.7, 27.5) | 26.0 (24.0, 28.3) | 21.6 (20.0, 23.3) | 22.6 (21.1, 23.8) | <0.001 |
| Heart rate, bpm | 68 (62, 75) | 67 (61, 73) | 69 (63, 76) | 70 (62, 74) | 71 (66, 80) | <0.001 |
| Systolic blood pressure, mmHg | 130 (120, 140) | 130 (120, 140) | 130 (120, 140) | 100 (118, 133) | 120 (110, 130) | <0.001 |
| Diastolic blood pressure, mmHg | 76 (70, 81) | 78 (70, 82) | 74 (69, 80) | 71 (66, 77) | 68 (64, 73) | <0.001 |
| Pulse pressure, mmHg | 50 (44, 62) | 50 (44, 61) | 52 (44, 64) | 51 (44, 60) | 50 (41, 64) | 0.383 |
| Family history of CAD, n (%) | 554 (31.5) | 335 (31.9) | 192 (33.5) | 6 (13.0) | 21 (28.8) | 0.035 |
| CV risk factors and prior CV disease |  |  |  |  |  |  |
| SMuRFs |  |  |  |  |  |  |
| Hypertension, n (%) | 1112 (63.8) | 377 (64.4) | 373 (65.1) | 16 (34.8) | 46 (63.0) | <0.001 |
| Diabetes, n (%) | 805 (46.2) | 474 (45.1) | 278 (48.5) | 16 (34.8) | 37 (50.7) | 0.191 |
| Hypercholesterolaemia, n (%) | 1396 (80.1) | 809 (77.0) | 494 (86.2) | 32 (69.6) | 61 (83.6) | <0.001 |
| Smoking, n (%) | 1025 (58.8) | 617 (58.7) | 345 (60.2) | 24 (52.2) | 39 (53.4) | 0.539 |
| SMuRFs-less, n (%) | 32 (1.8) | 14 (1.3) | 13 (2.3) | 2 (4.3) | 3 (4.1) | 0.128 |
| Peripheral artery disease, n (%) | 183 (10.5) | 91 (8.7) | 78 (13.6) | 4 (8.7) | 10 (13.7) | 0.013 |
| Old myocardial infarction, n (%) | 333 (19.1) | 201 (19.1) | 105 (18.3) | 12 (26.1) | 15 (20.5) | 0.621 |
| Prior PCI, n (%) | 343 (19.7) | 223 (21.2) | 100 (17.5) | 13 (28.3) | 7 (9.6) | 0.017 |
| Prior Cardiac arrest, n (%) | 2 (0.1) | 0 (0) | 2 (0.3) | 0 (0) | 0 (0) | 0.240 |
| Atrial fibrillation/flutter, n (%) | 74 (4.2) | 45 (4.3) | 24 (4.2) | 2 (4.3) | 3 (4.1) | 1.000 |
| Prior stroke, n (%) | 102 (5.9) | 53 (5.0) | 44 (7.7) | 3 (6.5) | 2 (2.7) | 0.103 |
| Comorbidity |  |  |  |  |  |  |
| Anemia, n (%) | 57 (3.3) | 17 (1.6) | 25 (4.4) | 3 (6.5) | 12 (16.4) | <0.001 |
| COPD, n (%) | 26 (1.5) | 12 (1.1) | 12 (2.1) | 0 (0) | 2 (2.7) | 0.273 |
| Cancer history, n (%) | 13 (0.7) | 8 (0.8) | 4 (0.7) | 0 (0) | 1 (1.4) | 1.000 |
| ACS presentation |  |  |  |  |  |  |
| Type of ACS, n (%) |  |  |  |  |  |  |
| STEMI | 229 (13.1) | 90 (8.6) | 110 (19.2) | 4 (8.7) | 25 (34.2) | <0.001 |
| NSTEMI | 227 (13.0) | 105 (10.0) | 99 (17.3) | 8 (17.4) | 15 (20.5) | <0.001 |
| Unstable angina | 1287 (73.8) | 856 (81.4) | 364 (63.5) | 34 (73.9) | 33 (45.2) | <0.001 |
| Killip class >1, n (%) | 67 (3.8) | 22 (2.1) | 26 (4.5) | 4 (8.7) | 15 (20.5) | <0.001 |
| The GRACE risk score, points | 98 (82, 114) | 94 (92, 108) | 99 (82, 119) | 105 (91, 118) | 120 (102, 155) | <0.001 |
| High risk at six months, n (%) | 356 (20.4) | 158 (15.0) | 150 (26.2) | 11 (23.9) | 37 (50.7) | <0.001 |
| Laboratory data and Echocardiographic data |  |  |  |  |  |  |
| White blood cell, 10<sup>9</sup>/L | 6.3 (6.3, 7.5) | 6.0 (5.2, 7.0) | 7.1 (6.0, 8.1) | 6.1 (5.0, 7.0) | 7.3 (6.2, 9.6) | <0.001 |
| Neutrophil-lymphocyte ratio | 2.2 (1.7, 3.0) | 1.0 (0, 1.0) | 2.4 (1.9, 3.2) | 2.3 (1.9, 3.1) | 3.1 (2.3, 4.2) | <0.001 |
| High-sensitivity C-reactive protein, mg/L | 1.7 (0.7, 3.5) | 0.78 (0.4, 1.3) | 4.6 (3.1, 8.9) | 0.7 (0.4, 1.3) | 9.5 (4.8, 18.5) | <0.001 |
| Triglycerides, mg/dL | 1.5 (1.0, 2.1) | 1.4 (1.0, 2.0) | 1.6 (1.2, 2.2) | 1.1 (0.7, 1.7) | 1.3 (1.0, 1.8) | <0.001 |
| Total cholesterol, mg/dL | 4.0 (3.4, 4.8) | 4.0 (3.4, 4.7) | 4.2 (3.6, 5.0) | 3.6 (3.1, 4.2) | 3.7 (3.4, 4.5) | <0.001 |
| LDL-C, mg/dL | 2.3 (1.8, 3.0) | 2.3 (1.8, 2.8) | 2.6 (2.0, 3.0) | 1.9 (1.6, 2.4) | 2.2 (1.8, 3.0) | <0.001 |
| HDL-C, mg/dL | 1.0 (0.9, 1.2) | 1.0 (0.9, 1.2) | 1.0 (0.8, 1.1) | 1.0 (0.9, 1.3) | 0.9 (0.8, 1.1) | <0.001 |
| B-type natriuretic peptide, pg/ml | 37 (21, 86) | 33 (20, 67) | 33 (20, 67) | 40 (24, 90) | 197 (52, 197) | <0.001 |
| Albumin, g/L | 42 (40, 45) | 43 (40, 45) | 45 (23, 118) | 37 (36, 39) | 36 (34, 38) | <0.001 |
| Creatinine clearance rate, ml/min | 96.7 (78.8, 116.7) | 98.1 (81.2, 117.6) | 97.8 (78.5, 120.0) | 83.4 (67.1, 97.5) | 80.1 (66.1, 92.9) | <0.001 |
| Uric acid, µmol/L | 333.2 (289.3, 397.8) | 324.5 (287.0, 390.2) | 345.3 (298.3, 413.2) | 297.7 (248.0, 365.3) | 325.4 (296.7, 398.2) | <0.001 |
| Fasting plasma glucose, mmol/L | 5.8 (5.2, 7.0) | 5.8 (5.2, 6.8) | 6.0 (5.3, 7.2) | 5.6 (4.9, 6.7) | 6.1 (5.3, 7.4) | 0.007 |
| Glycated hemoglobin A1c, % | 6.1 (5.6, 7.1) | 6.0 (5.5, 7.0) | 6.3 (5.6, 7.3) | 6.0 (5.5, 6.5) | 6.3 (5.7, 7.4) | 0.002 |
| LVSD (EF < 50%), n (%) | 106 (6.1) | 42 (4.0) | 45 (7.9) | 6 (13.0) | 13 (17.8) | <0.001 |
| Angiographic data |  |  |  |  |  |  |
| Left main coronary artery lesion, n (%) | 136 (7.8) | 71 (6.8) | 55 (9.6) | 4 (8.7) | 6 (8.2) | 0.213 |
| Multivessel coronary artery disease, n (%) | 1457 (83.6) | 866 (82.4) | 493 (86.0) | 35 (76.1) | 63 (86.3) | 0.116 |
| Chronic total occlusion, n (%) | 367 (21.1) | 208 (19.8) | 141 (24.6) | 5 (10.9) | 13 (17.8) | 0.036 |
| SYNTAX score | 20 (13, 28) | 19 (12, 27) | 22 (15, 29) | 21 (11, 30) | 26 (17, 34) | <0.001 |
| Complete revascularization, n (%) | 1069 (61.3) | 687 (65.4) | 312 (54.5) | 31 (67.4) | 39 (53.4) | <0.001 |
| Percutaneous transfemoral route, n (%) | 27 (1.5) | 21 (2.0) | 4 (0.7) | 0 (0) | 2 (2.7) | 0.122 |
| Medical therapy |  |  |  |  |  |  |
| Pre-admission, n (%) |  |  |  |  |  |  |
| Antiplatelet agents | 1272 (73.0) | 781 (74.3) | 403 (70.3) | 37 (80.4) | 51 (69.9) | 0.205 |
| Statin | 1252 (71.8) | 772 (73.5) | 389 (67.9) | 39 (84.8) | 52 (71.2) | 0.021 |
| Discharge, n (%) |  |  |  |  |  |  |
| Dual antiplatelet therapy | 1727 (99.1) | 1047 (99.6) | 565 (98.6) | 45 (97.8) | 70 (95.9) | 0.013 |
| Aspirin plus clopidogrel | 1587 (91.0) | 968 (92.1) | 508 (88.7) | 42 (91.3) | 69 (94.5) | 0.086 |
| Aspirin plus ticagrelor | 140 (8.0) | 79 (7.5) | 57 (9.9) | 3 (6.5) | 1 (1.4) | 0.050 |
| Statin | 1743 (100.0) | 1051 (100.0) | 573 (100.0) | 46 (100.0) | 73 (100.0) | - |
| Beta-blockers | 1225 (70.3) | 728 (45.8) | 418 (72.9) | 28 (60.9) | 51 (69.9) | 0.212 |
| ACEI/ARB | 841 (48.3) | 481 (45.8) | 301 (52.5) | 20 (43.5) | 39 (53.4) | 0.045 |
| Furosemide | 114 (6.5) | 53 (5.0) | 46 (8.0) | 2 (4.3) | 13 (17.8) | <0.001 |
| Malnutrition classified by NRI |  |  |  |  |  |  |
| Moderate or severe malnutrition, n (%) | 37 (2.1) | 0 (0) | 0 (0) | 14 (30.4) | 23 (31.5) | <0.001 |
| Outcomes |  |  |  |  |  |  |
| MACEs, n (%) | 351 (20.1) | 170 (16.2) | 144 (25.1) | 7 (15.2) | 30 (41.1) | <0.001 |
| Cardiac death, n (%) | 37 (2.1) | 12 (1.1) | 14 (2.5) | 2 (4.3) | 9 (12.3) | <0.001 |
| Non-fatal MI, n (%) | 43 (2.5) | 20 (1.9) | 19 (3.3) | 1 (2.2) | 3 (4.1) | 0.268 |
| Non-fatal stroke, n (%) | 22 (1.3) | 9 (0.9) | 12 (2.1) | 1 (2.2) | 0 (0) | 0.121 |
| Unplanned revascularization, n (%) | 249 (14.3) | 129 (12.3) | 99 (17.3) | 3 (6.5) | 18 (24.7) | 0.003 |

a. Hs-CRP level over the cut-off value of 2.2mg/L was classified as elevated CRP.
Abbreviations: ACEI, angiotensin-converting enzyme inhibitor; ACS, acute coronary syndrome; ARB, angiotensin receptor blockade; CAD, coronary artery disease; COPD, chronic obstructive pulmonary disease; CV, cardiovascular; EF, ejection fraction; GRACE, the Global Registry of Acute Coronary Events; HDL-C, high-density lipoprotein cholesterol; LDL-C, low-density lipoprotein cholesterol; LVSD, left ventricular systolic dysfunction; MACEs, major adverse cardiac events; NRI, Nutrition risk index; NSTEMI, non-ST-elevation myocardial infarction; PAD, peripheral artery disease; PCI, percutaneous coronary intervention; STEMI, ST-elevation myocardial infarction; TC, total cholesterol; TG, triglycerides; WBC, white blood cell.

**Table 2.**
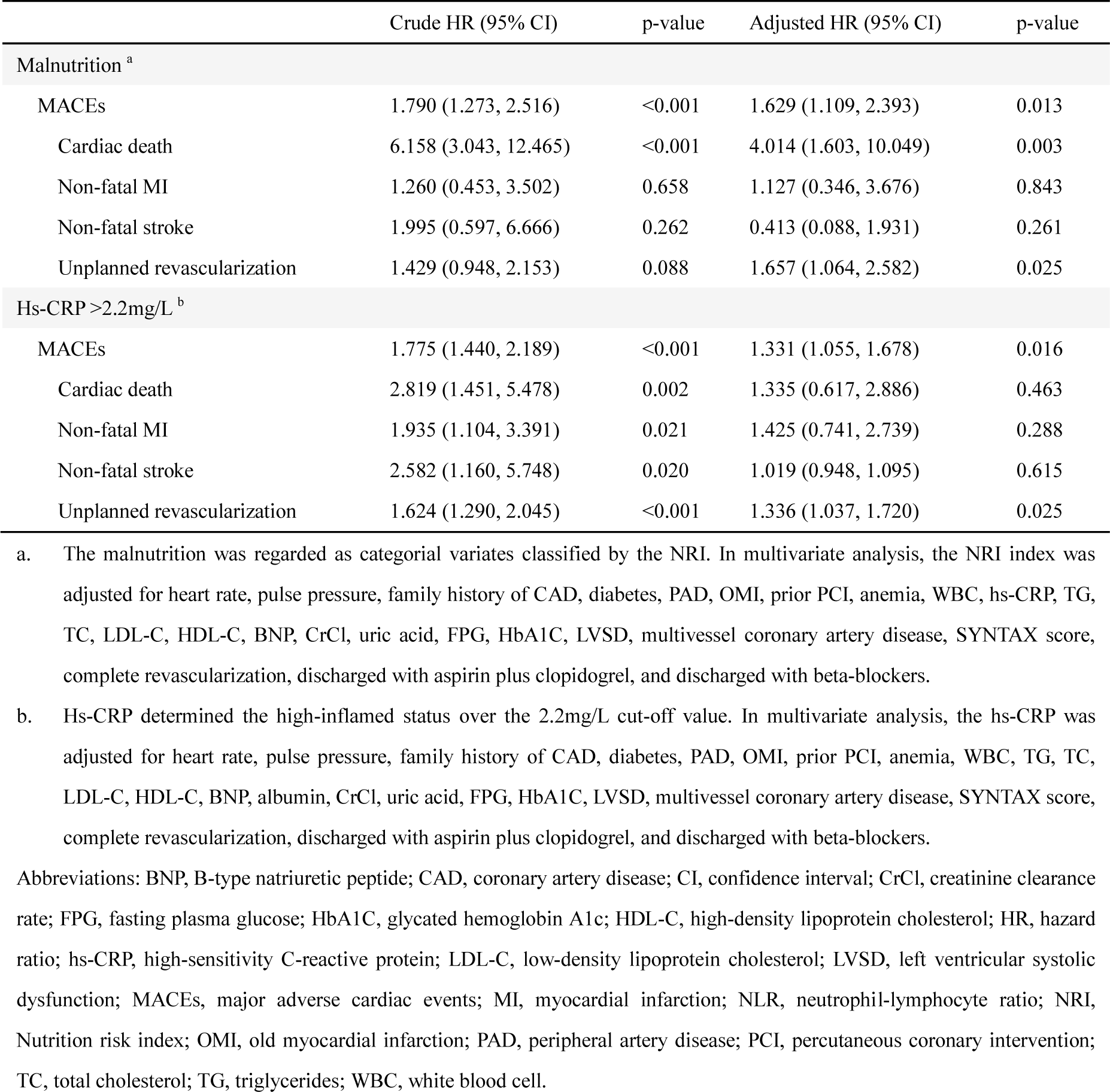
Predictive value of malnutrition or elevated hs-CRP for outcomes in univariate and multivariate proportional.

**Figure 1.**
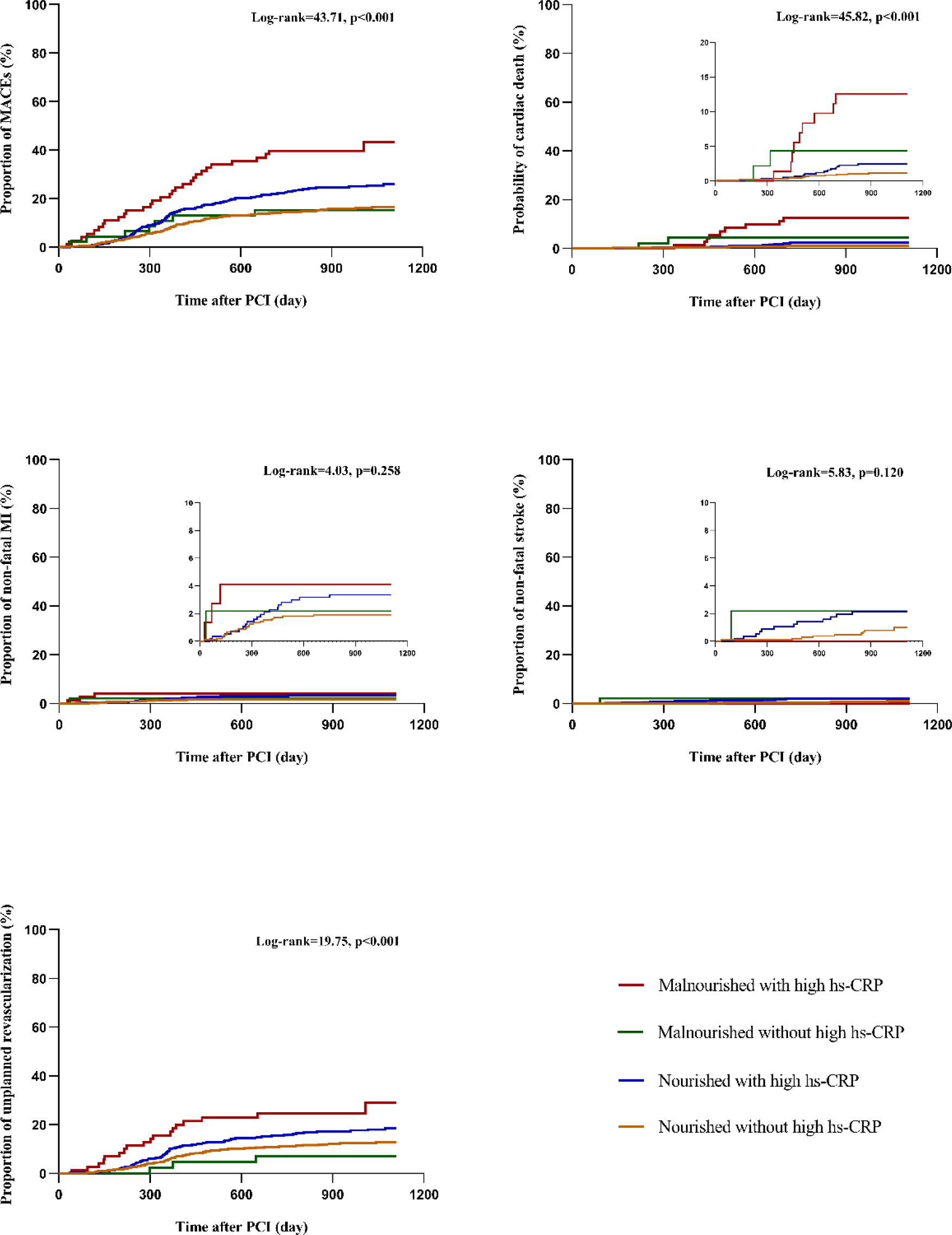
Kaplan-Meier survival curves for MACEs and each component up to 3 years.

With nourished without the elevated hs-CRP group as the reference, malnourished patients with elevated hs-CRP showed significantly higher MACEs, cardiac death, and unplanned revascularization in multivariate analyses (all p <0.05); the nourished with elevated hs-CRP group presented with obviously increased cardiac death and unplanned revascularization (all p <0.05).

Interestingly, patients in the malnourished group without elevated hs-CRP exhibited no significant difference in affecting MACEs and each component compared with the reference group (**Table 3**). The same analyses were done in patients classified by hs-CRP and the COUNT score, which showed no significant differences among the groups **(Supplementary Table 4)**.

**Table 3.**
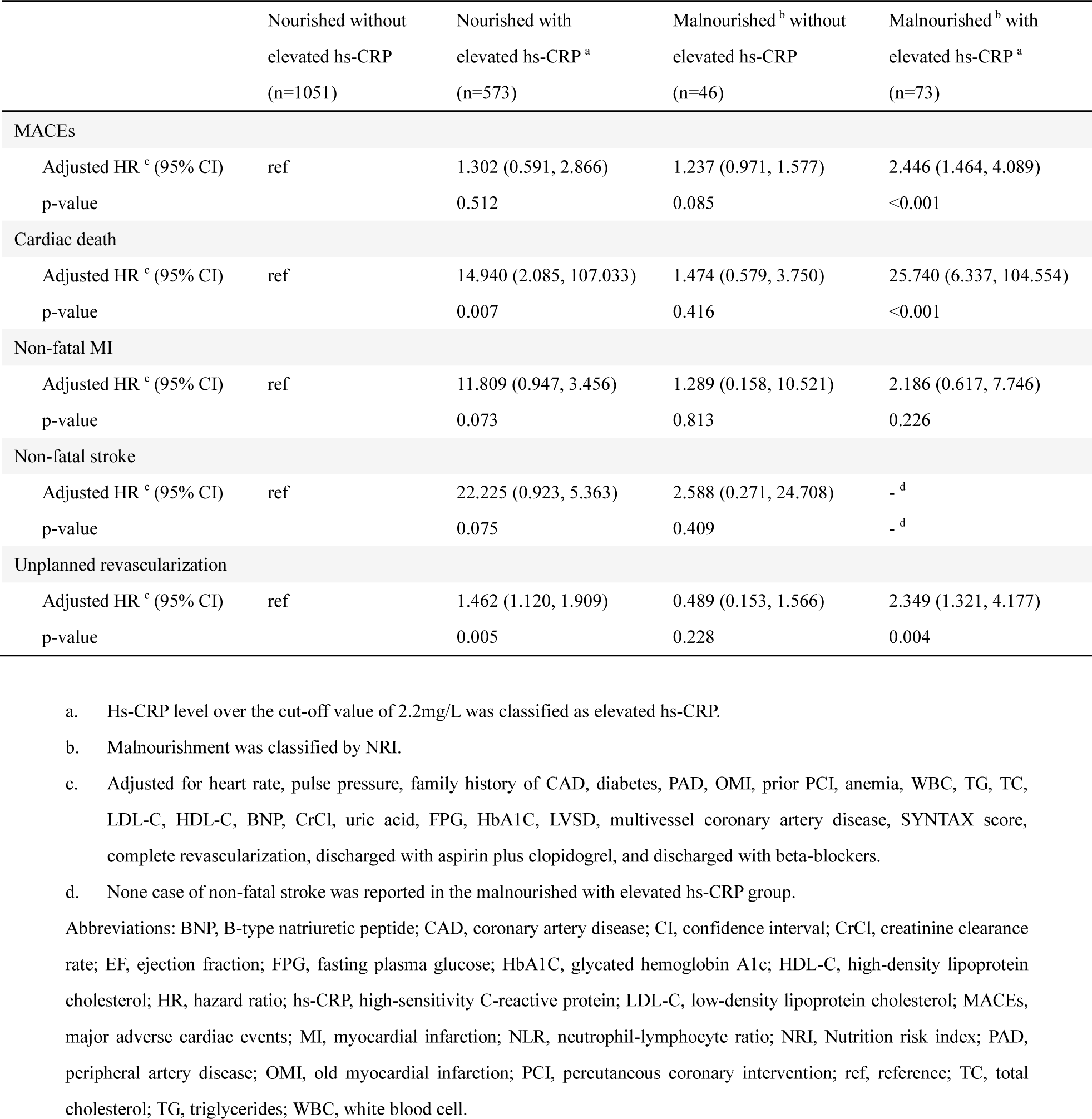
Predictive value of each group on outcomes in multivariate proportional hazards regression analyses.

To further understand whether the effect of elevated hs-CRP or malnutrition on the MACEs noted in the whole cohort was influenced by baseline nutritional or inflammatory status, we conducted stratified analyses within subgroups and calculated interaction statistics. As shown in **Table 4A**, the p-value for interaction between nutritional status (nourished vs. malnourished) and hs-CRP levels (≤2.2mg/L vs. >2.2mg/L) on MACEs was 0.248; when including hs-CRP in the model as a continuous variable, it remained a risk enhancer among the malnourished (p=0.003) but not the nourished (p = 0.620), with evidence for interaction (p for interaction =0.001). Simultaneously, as it was included in the model as a continuous variable, the NRI score still shared a significant correlation with MACEs in high-inflamed patients (p=0.003) while failing to present it in those without (p=0.869), with evidence for interaction (p for interaction =0.005) (**Table 4B**). When hs-CRP and malnutrition were both included in the model as continuous variables, there was evidence for effect modification between each other on the association of MACEs (P for interaction = 0.046).

**Table 4.** Interplay of elevated hs-CRP or malnutrition on MACEs.

**A) Examining the effect of inflammation on MACEs in patients stratified by nutritional status.**
| Examined variable | Nourished subpopulation |  |  | Malnourished subpopulation |  |  | p for interaction |
| --- | --- | --- | --- | --- | --- | --- | --- |
|  | Events (%) | Adjusted HR <sup>a</sup> (95% CI) | p-value | Events (%) | Adjusted HR <sup>a</sup> (95% CI) | p-value |  |
| Hs-CRP >2.2mg/L | 324 (20.0) | 1.237 (0.971, 1.576) | 0.085 | 37 (31.1) | 4.858 (1.353, 17.440) | 0.015 | 0.248 |
| Hs-CRP level, mg/L | 324 (20.0) | 1.006 (0.982, 1.030) | 0.620 | 37 (31.1) | 1.084 (1.027, 1.143) | 0.003 | 0.001 |

**B) Examining the effect of malnutrition on MACEs in patients stratified by inflammatory status.**
| Examined variable | Hs-CRP ≤2.2mg/L subpopulation |  |  | Hs-CRP >2.2mg/L subpopulation |  |  | p for interaction |
| --- | --- | --- | --- | --- | --- | --- | --- |
|  | Events (%) | Adjusted HR <sup>a</sup> (95% CI) | p-value | Events (%) | Adjusted HR <sup>a</sup> (95% CI) | p-value |  |
| Malnourished status | 177 (16.1) | 1.287 (0.587, 2.832) | 0.531 | 174 (26.9) | 1.927 (1.232, 3.014) | 0.004 | 0.248 |
| The NRI score <sup>b</sup> | 177 (16.1) | 1.002 (0.997, 1.028) | 0.869 | 174 (26.9) | 0.965 (0.943, 0.988) | 0.003 | 0.005 |

a. Adjusted for heart rate, pulse pressure, family history of CAD, diabetes, PAD, OMI, prior PCI, anemia, WBC, TG, TC, LDL-C, HDL-C, BNP, CrCl, uric acid, FPG, HbA1C, LVSD, multivessel coronary artery disease, SYNTAX score, complete revascularization, discharged with aspirin plus clopidogrel, and discharged with beta-blockers.
b. The NRI score was calculated using the formulas $[1.519 \times \text{serum albumin (g/L)}] + 41.7 \times (\text{current weight/ideal weight})$ for patients under 65 years old and $[1.489 \times \text{serum albumin (g/L)}] + 41.7 \times (\text{current weight/ideal weight})$ for those over 65 years old. A higher NRI score suggests better nutritional status.
Abbreviations: BNP, B-type natriuretic peptide; CAD, coronary artery disease; CI, confidence interval; CrCl, creatinine clearance rate; EF, ejection fraction; FPG, fasting plasma glucose; HbA1C, glycated hemoglobin A1c; HDL-C, high-density lipoprotein cholesterol; HR, hazard ratio; hs-CRP, high-sensitivity C-reactive protein; LDL-C, low-density lipoprotein cholesterol; MACEs, major adverse cardiac events; MI, myocardial infarction; NLR, neutrophil-lymphocyte ratio; NRI, Nutrition risk index; OMI, old myocardial infarction; PAD, peripheral artery disease; PCI, percutaneous coronary intervention; ref, reference; TC, total cholesterol; TG, triglycerides; WBC, white blood cell.

Finally, the incremental predictive effect of hs-CRP or NRI beyond the established GRACE risk model was evaluated. As adding each variate into the GRACE model significantly increased the original predictive value on MACEs or cardiac death, a synergistic effect showed when combining both hs-CRP and NRI with the GRACE model (**Table 5**).

**Table 5.** Incremental effect of inflammation or malnutrition beyond the GRACE risk model to predict cardiac death.

| Model | C-index (95%CI) | p-value | Net reclassification improvement (95%CI) | p-value | Integrated discrimination Improvement (95%CI) | p-value |
| --- | --- | --- | --- | --- | --- | --- |
| MACEs |  |  |  |  |  |  |
| GRACE <sup>a</sup> | 0.538 (0.514, 0.562) | 0.003 | ref |  | ref |  |
| GRACE + NRI <sup>b</sup> | 0.554 (0.530, 0.577) | <0.001 | 0.128 (0.011, 0.245) | 0.032 | 0.005 (0.002, 0.009) | 0.004 |
| GRACE + hs-CRP <sup>c</sup> | 0.590 (0.566, 0.613) | <0.001 | 0.196 (0.088, 0.303) | <0.001 | 0.006 (0.002, 0.010) | 0.005 |
| GRACE + NRI + hs-CRP | 0.594 (0.570, 0.617) | <0.001 | 0.210 (0.094, 0.326) | <0.001 | 0.010 (0.005, 0.016) | <0.001 |
| Cardiac death |  |  |  |  |  |  |
| GRACE <sup>a</sup> | 0.699 (0.677, 0.721) | <0.001 | ref |  | ref |  |
| GRACE + NRI <sup>b</sup> | 0.733 (0.712, 0.754) | <0.001 | 0.629 (0.318, 0.940) | <0.001 | 0.020 (0.007, 0.033) | 0.002 |
| GRACE + hs-CRP <sup>c</sup> | 0.702 (0.680, 0.723) | <0.001 | 0.369 (0.050, 0.688) | 0.023 | 0.006 (0.001, 0.012) | 0.049 |
| GRACE + NRI + hs-CRP | 0.738 (0.717, 0.759) | <0.001 | 0.666 (0.361, 0.971) | <0.001 | 0.023 (0.009, 0.037) | 0.002 |

- a. The eight variables that constitute the GRACE risk score are age, history of heart failure, history of acute MI, heart rate and systolic blood pressure at admission, ST-segment depression, serum creatinine at admission, and elevated myocardial necrosis markers or enzymes. - b. NRI was introduced to the GRACE model as a categorical variate. - c. Hs-CRP was introduced to the GRACE model as a categorical variate.
Abbreviations: C-index, concordance index; CI, confidence interval; GRACE, the Global Registry of Acute Coronary Events; hs-CRP, high-sensitivity C-reactive protein; MACEs, major adverse cardiac events; NRI, Nutrition risk index; ref, reference.

## Discussion

This study of retrospective data is the first to highlight the importance of an easily overlooked subgroup of ACS patients, who suffered a double burden of malnutrition and high-inflamed status. We observed that malnutrition combined with elevated hs-CRP is significantly associated with an increased risk of long-term cardiac outcomes among ACS patients, and inflammation and malnutrition may interplay by amplifying the harmful effects of each other.

The prevalence of malnutrition identified throughout the present ACS cohort and previous studies varied across different screening tools, population characteristics, and sample sizes. Komici et al. evaluated the nutritional status of 174 elderly AMI patients by Mini Nutritional Assessment (MNA), and 12% of patients were malnourished^23^; Tonet et al. reported that 4% of elderly ACS patients were undernourished using the MNA–Short Form^24^. Li et al. surveyed 5,710 CAD patients with prediabetes or diabetes (49% were ACS), showing that malnutrition rate was 10%, 8%, 46%, and 20% according to GLIM criteria, PNI, COUNT score, and NRI, respectively^25^. In another cohort with 705 patients diagnosed with ACS and chronic kidney disease (CKD), malnutrition was found in 30%, 80%, and 90% of the population according to the PNI, CONUT, and GNRI, respectively^26^. Data from 5,062 ACS patients suggested that malnutrition prevailed in over 50% of cases based on the CONUT score, NRI, or PNI^27^. Another large cohort involving 10,161 AMI patients who underwent PCI showed that 11% were at risk of malnutrition as determined by GNRI^28^. Recently, Kong et al. observed 1,829 AMI patients, 76.5%, 61.8%, and 39.3% of whom were malnourished according to the CONUT score, NRI, and PNI, respectively^9^. These variations reflect the necessity and importance of setting universal, standardized, and pragmatic nutritional evaluation criteria to facilitate risk stratification and management. In 2018, the American Society for Parenteral and Enteral Nutrition (ASPEN) led the launching of the Global Leadership Initiative on Malnutrition (GLIM) as a consensus-based criterion of malnutrition^29^; however, the need to be equipped with body composition measuring techniques may hinder its wide use in a general setting, and it has yet to get prospective clinical validation among CVD. Although the COUNT score has been frequently used in recent studies among CVD patients, its scoring outcome could be strongly perturbed by lipid-lowering therapies, especially in those with past pertinent history and are potentially at high risk, which partly explains why the COUNT score lacked discrimination ability in our cohort with 72% of the participants received statin pre-admission. Besides, the PNI showed a severely poor sensitivity in our cohort, which was in line with some previous studies ^9,25^. The NRI displayed the best risk-discriminating ability among the three screening tools in our cohort, which is inconsistent with an earlier study suggesting that the CONUT score had a higher predictive ability in AMI patients than the NRI and PNI. Thus, studies based on large ACS populations and thorough assessments are needed to establish a consensus on which nutritional evaluation method to use for patients with ACS.

In our cohort, elders, females, and lean people were found more frequently in the malnourished group, and they also displayed lower cholesterol levels; these clinical characteristics are consistent with the former studies ^9,27^. One of the notable observations of our study is that a heavy burden of systemic inflammation marked by elevated hs-CRP prevailed in malnourished ACS patients by over 60%, suggesting a possible close interaction between malnutrition and inflammation in the pathophysiology and progression of CVD. Inflammation is thought to be a key driver for sarcopenia elicited by an underlying illness, leading to disease-related malnutrition and increased mortality^30,31^. CAD has been recognized as not only a lipid metabolic disorder but also an inflammation-mediated systemic disease, with the pro- and anti-inflammatory reactions interacting to exacerbate or ameliorate the atherosclerotic process and causing catabolic activity, energy expenditure and function loss. One of the primary underlying mechanisms is the circulating cytokines released from the systemic inflammatory response (i.e., interleukin 6, interleukin 1β, and tumor necrosis factors), triggering mechanisms that contribute to the pathogenesis of malnutrition, including brain circuits disorder, protein degradation, and neuroendocrine disturbance, ultimately causing disease-related anorexia, sarcopenia, cognitive decline, and frailty^2,3,32,33^. In 2012, ASPEN and the Academy of Nutrition and Dietetics launched a consensus statement emphasizing the need to identify inflammation early in the diagnostic procedure of malnutrition to determine its etiology and guide treatment^34^. In a traumatic and acute stress-causing event like ACS, albumin serves as a practical plasma biomarker that can be used to indicate and monitor catabolic metabolism and systemic inflammatory response to reflect disease severity; hypoalbuminemia was also one of the most striking features of patients with MACEs in our cohort.

Nutritional status influences the body’s inflammatory response, and consistent nutritional management is crucial for patients with critical illness to recover from inflammatory injury ^33,35^. Although the prevalence of malnutrition exhibited significant discrepancies, its association with poorer outcomes remains clear and sound ^23–28^. However, the importance of screening and monitoring for malnutrition in patients with ACS hasn’t drawn enough concern from clinical guidelines and practice^13,14^. The interaction analyses of our study suggested the bidirectional association between inflammation and malnutrition, while the present stratification standards to define high inflammation or malnutrition seemingly affected the results. In this sense, studies with larger sample sizes and ACS-suitable criteria for stratification seemed indispensable to facilitate the understanding of the interplay between inflammation and malnutrition. Given the existing results, we could speculate that patients with worse nutritional conditions may urgently need deactivating inflammation, and patients with higher hs-CRP would benefit more from nutritional support. Previous studies among hospitalized patients showed that baseline inflammatory status was associated with their response to nutritional therapy and that nutritional support alone is inadequate to prevent muscle loss during severe, sustained, or repeated bouts of inflammation^36,37^, which emphasizes the importance of considering the double burden of inflammation and malnutrition in disease assessment, intervention, and monitoring. Regarding ACS patients, there is still a lack of evidence investigating the treatment response to nutritional support or anti-inflammatory therapy among subgroups with different degrees of inflammation or malnutrition.

To the best of our knowledge, the present study is the first analysis to investigate the interplay between inflammation and malnutrition in affecting cardiac outcomes among ACS patients, which would provide preliminary implications for future research. We are also aware of several limitations. First, as a retrospective observational study, we couldn’t conclude direct causality; we also may not accounted for other potential confounding effects and unknown clinical or subclinical illness. Second, although we tested and compared the predictive ability of the three malnutrition screening tools, their diagnostic accuracy or practical value among ACS patients was yet to be confirmed. This is also a common problem in similar studies that need to be solved. Third, the serum hs-CRP, albumin, and body weight were measured only once at admission and are subject to measurement error and physiologic variability; the dynamic and long-term monitoring of inflammatory and nutritional status changes would provide valuable and indispensable information. Fourth, as it was a single-center study based on Chinese people, the results should be carefully interpreted and extended to other ethnicities.

## Conclusions

Among patients with ACS undergoing PCI, the double burden of inflammation and malnutrition was not uncommon and strongly associated with long-term cardiac outcomes in a synergistic manner. Identifying patients’ inflammatory and nutritional status will better facilitate the modern individualized care of ACS. Future prospective studies are needed to evaluate whether the heterogeneity in the therapeutic effect of nutritional support or anti-inflammatory interventions exists in patients with different inflammatory or nutritional burdens.

## Funding

This research was funded by the Beijing Municipal Natural Science Foundation, General Program (7232039), and the Capital Health Research and Development of Special (2022-2-1052).

## Data Availability

The datesets uesd and/or analyzed during the current study are avalible from the corresponding author on reasonable request.

## Notes

### Competing Interest Statement

The authors have declared no competing interest.

### Author Declarations

This study followed the Helsinki Declaration of Human Rights and was approved by the institutional review board (IRB) of Beijing Anzhen Hospital (IRB number: 2016034x). The IRB waived the need for written patient consent as this study involved a retrospective analysis of clinically acquired data.

